# Bladder Cancer Prognosis Using Deep Neural Networks and Histopathology Images

**DOI:** 10.1101/2022.03.04.22271918

**Authors:** Wayner Barrios, Behnaz Abdollahi, Manu Goyal, Qingyuan Song, Matthew Suriawinata, Ryland Richards, Bing Ren, Alan Schned, John Seigne, Margaret Karagas, Saeed Hassanpour

## Abstract

Recent studies indicate bladder cancer is among the top 10 most common cancer in the world [1]. Bladder cancer frequently reoccurs, and prognostic judgments may vary among clinicians. Classification of histopathology slides is essential for accurate prognosis and effective treatment of bladder cancer patients, as a favorable prognosis might help to inform less aggressive treatment plans. Developing automated and accurate histopathology image analysis methods can help pathologists in determining the prognosis of bladder cancer. In this study, we introduced Bladder4Net, a deep learning pipeline to classify whole-slide histopathology images of bladder cancer into two classes: low-risk (combination of PUNLMP and low-grade tumors) and high-risk (combination of high-grade and invasive tumors). This pipeline consists of 4 convolutional neural network (CNN) based classifiers to address the difficulties of identifying PUNLMP and invasive classes. We evaluated our pipeline on 182 independent whole-slide images from the New Hampshire Bladder Cancer Study (NHBCS) [22] [23] [24] collected from 1994 to 2004 and 378 external digitized slides from The Cancer Genome Atlas (TCGA) database [26]. The weighted average F1-score of our approach was 0.91 (95% confidence interval (CI): 0.86–0.94) on the NHBCS dataset and 0.99 (95% CI: 0.97–1.00) on the TCGA dataset. Additionally, we computed Kaplan-Meier survival curves for patients predicted as high-risk versus those predicted as low-risk. For the NHBCS test set, patients predicted as high-risk had worse overall survival than those predicted as low-risk, with a Log-rank P-value of 0.004. If validated through prospective trials, our model could be used in clinical settings to improve patient care.

## Introduction

Recent studies indicate bladder cancer is among the top 10 most common cancer in the world [1]. Most bladder cancer cases are urothelial carcinoma. Approximately 75-85% of patients with bladder cancer are classified as non-muscle-invasive bladder cancer (NMIBC). Furthermore, around 50% of NMIBC patients experience one or more disease recurrences, and the treatment procedure is different from patients diagnosed with muscle-invasive bladder cancer (MIBC) [2]. Urothelial carcinomas are graded according to the degree of tumor cellular and architectural atypia. The cancer grade has an important role in deciding the treatment plan, so if not determined accurately, the patient might undergo unnecessary treatments. The World Health Organization (WHO) 1973 and World Health Organization/International Society of Urological Pathology (WHO/ISUP) classifications are widely used for tumor grading, but these methods have relatively high intra- and inter-observer variabilities [3] [4]. Several studies compared different grading systems and their effect on choosing the best treatment [5] [6] [7] [8]. A study evaluated WHO 1973 classification from 11 pathologists, and inter-observer agreement was slight to moderate (κ = 0.19 − 0.44) [9]. Another study measured inter-observer agreement among 6 pathologists and showed that WHO/ISUP classification is slightly better than WHO 1973 [10]. Therefore, new methods should be sought to help pathologists diagnose bladder cancer.

Stage and grade of bladder tumors are important criteria in cancer treatment. The cancer stage consists of the location of the cancer cells and how far they have grown. Higher stages indicate whether the tumor has grown away from the surface. Urothelial carcinoma pathologic stages are named as Ta (Papillary tumor without invasion), TIS (Carcinoma in situ (CIS)), T1(tumor invades the connective tissue under the surface lining), T2 (tumor invades the muscle layer), T3 (tumor invades perivesical soft tissue), and T4 (extravesical tumor directly invades into other organs or structures). According to WHO 2016 classification, NMIBC is divided into three groups: Ta, Tis, and T1, while T2, T3, and T4 are MIBC. In cases of low grade, the cancer cells show morphology with less atypia and more resembling normal urothelial cells and grow slowly. Clinical research has demonstrated that the most common bladder tumors are low-grade [11]. On the other hand, high-grade cancer cells show more irregular and atypical morphology and can be found in both NMIBC and MIBC. It is essential to accurately differentiate between low and high grades because different treatments are available for various grade tumors. For example, high-grade cells in NMIBC need prompt treatment to avoid the spread of cancer.

Papillary urothelial neoplasm of low malignant potential (PUNLMP) was first introduced by the WHO/ISUP in 1998 as a new entity of bladder cancer [12]. PUNLMP and low-grade urothelial carcinoma are two bladder cancer types that are not easily distinguishable based on cell morphology under the microscope. Because of similarities between the two cancer types, the pathologic diagnostic accuracy for their differentiation is about 50% [13]. While the distinction between PUNLMP and low-grade is deemed essential by some pathologists, recent studies showed that separating PUNLMP and low-grade is not clinically crucial [5]. Of note, high-grade tumor cells are found in various tumor stages, and MIBC cancer type is considered high-grade.

Histological classification of bladder cancer has significant implications in the prognosis and treatment of patients. Moreover, detecting and classifying histologic patterns such as PUNLMP and low-grade urothelial carcinoma under the microscope is a time-consuming and challenging task for pathologists. Manual classification of bladder cancer histological patterns has a high error rate due to the similarity of histological features. Therefore, clinical information such as the cancer stage is commonly used for a more accurate prognosis. Automated image analysis using deep learning techniques can assist pathologists in providing faster and more consistent results. Additionally, these techniques can be improved by providing new data and associated annotated labels by several pathologists so that the model can be trained based on the expert opinion of multiple pathologists. In recent years, deep learning models, such as convolutional neural networks (CNNs), have been applied to a variety of computer vision tasks as well as biomedical applications [14] [15] [16]. CNN-based models have shown great promise in learning morphological characteristics of different cancer types from histological images [17] [18] [19] [20] [21].

Automated image analysis methods to classify and visualize various cancer patterns in high-resolution whole-slide images can help pathologists avoid errors and reduce their assessment time. In this study, we introduced a CNN-based model for the classification of urothelial bladder cancer based on whole-slide histopathology images to distinguish between low-risk and high-risk groups where low-risk class includes PUNLMP and low-grade cases, and the high-risk class includes high-grade and invasive cases.

## Materials and methods

### Datasets

For the model development and evaluation, we used images from the New Hampshire Bladder Cancer Study or NHBCS [22] [23] [24]. Risk factors for bladder cancer have been widely explored in previous reports from this study [25]. For external evaluation, we utilized histology images from The Cancer Genome Atlas (TCGA) [26]. The details of these datasets are included below.

### New Hampshire Bladder Cancer Study (NHBCS) Dataset

This dataset contains 838 whole-slide images from 1994 to 2004 as part of the NHBCS [27]. These hematoxylin and eosin (H&E) stained surgical resection slides were digitized by Aperio AT2 scanners (Leica Biosystems, Wetzlar, Germany) at 20×□magnification (0.50 μm/pixel).

### The Cancer Genome Atlas (TCGA) Dataset

We collected 378 whole-slide images from TCGA for external validation. The distribution of these whole-slide images used in this study is summarized in Table 1.

**Table 1.** Distribution of collected whole-slide images from four classes (PUNLMP, low-grade, high-grade, IUC) and distribution of low-risk and high-risk images in our datasets.

| **Histologic Subtype** | **Internal (NHBCS) training set** | **Internal (NHBCS) test set** | **External (TCGA) test set** |
| --- | --- | --- | --- |
| Low-risk<br>(PUNLMP + Low Grade) | 248<br>(94 + 154) | 107<br>(39 + 68) | 11<br>(11 + 0) |
| High-risk<br>(High Grade + IUC) | 177<br>(144 + 33) | 75<br>(62 + 13) | 367<br>(0 + 367) |
| Total | 425 | 182 | 378 |

### Data Annotation

The tumor histologic subtypes in NHBCS dataset were confirmed independently by two expert pathologists (A.S. & B.R.) from the Department of Pathology and Laboratory Medicine at Dartmouth–Hitchcock Medical Center (DHMC) based on a standard histopathology review. In NHBCS dataset, 637 whole-slides images were categorized into Papillary Urothelial Neoplasm of Low Malignant Potential (PUNLMP), Low-Grade Papillary Urothelial Carcinoma (low-grade, noninvasive), High-Grade Papillary Urothelial Carcinoma (high-grade, noninvasive), and Invasive Urothelial Carcinoma (IUC). Among these slides, 34 were classified as Carcinoma in Situ (CIS). Because of the small number of available CIS cases, we removed them from our study. In addition, 31 cases were labeled as others excluded from our study. We used 607 whole-slides from four classes (PUNLMP, low-grade, high-grade, and IUC) in our analysis. We combined PUNLMP and low-grade whole-slide images into a single class because of their similarity as they are both noninvasive and low risk cancers. Also, high-grade and IUC are merged into one group because they are considered high-risk cancers. We established the ground truth labels for each whole-slide image in our NHBCS data sets based on the consensus opinion of the two pathologists. If there was any disagreement, an expert pathologist (B.R.) re-reviewed the whole-side image and resolved the disagreement. We randomly partitioned these slides into an internal training set of 425 slides (∼70% of the NHBCS dataset) and an internal test set of 182 slides (∼30% of the NHBCS dataset).

Two pathologists (R.R. & B.R.) manually annotated the whole-slide images in our internal NHBCS training set using the Automated Slide Analysis Platform (ASAP) [28]. Regions of interests in each whole-slide image in our training set are annotated with bounding boxes at the highest resolution for each image. The annotated areas are split into smaller patches for training a patch-level classifier. As noted above, the groud truth labels for whole-slide images in our internal NHBCS test set were based on independent classification of two pathologists (B.R. and A.S.). The labels for the external TCGA test set were established based on the provided metadata from the TCGA database and additional confirmation by our study’s expert pathologist (B.R.).

### Bladder4Net: Deep Learning Pipeline

In this study, we developed a deep learning-based model to distinguish between low- and high-risk bladder cancer cases where the low-risk class includes PUNLMP and low-grade cases, and the high-risk class includes high-grade and invasive cases. This deep learning pipeline, named Bladder4Net, is shown in Figure 1. We classify each patch in a whole-slide image with binary classifiers. The portion of patches classified as a subtype in a whole-slide image is included in a vector for all classes. Of note, the ratio of PUNLMP and low-grade patches are added to represent low-risk patches, and the ratio of high-grade and invasive are combined to represent high-risk patches. A Gaussian process classifier was trained on low-risk and high-risk patch ratios using the same training and test set partitioning used for training the CNN classifier. The details of this pipeline are included below.

**Figure 1.**
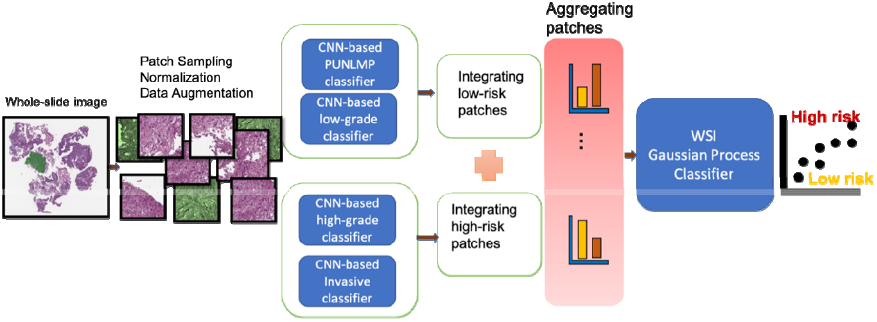
Overview of our classification pipeline. Tissue patches are extracted from whole-slide images using a sliding-window method with 1/3 overlap after background and marker removal. Deep neural networks extract histology features of the patches and classify them. The ratio of patches from each class is computed. We classify a whole-slide using a Gaussian process classifier based on the computed ratio of low-risk and high-risk classes.

### Patch classification

Analyzing large histology images using deep learning models requires substantial memory resources. Therefore, we split each whole-slide image into fixed-size patches (224×224 pixels) with 1/3 overlapping.The Bladder4Net pipeline consists of four binary ResNet-18 [29] deep learning models that operate at the patch level for each class. We randomly select 10% of whole-slides images in the training partition for hyperparameter tuning in order to find the best hyperparameters during training process. We selected patches in annotated areas for training and evaluating the patch-level classifiers. We normalized the color intensity of patches and used standard data augmentation methods, including random vertical and horizontal flips and color jittering, which its parameters selected based on the random subsampling of patches in each class. Our model was trained on 260,610 patches (average 613 patches per whole-image slide), including 107,379 high-risk and 153,231 low-risk patches. To address the class imbalance, we used a weighted random sampler method for generating the training batches. For model training, we trained a ResNet-18 [29] initialized using normal distribution initialization. All four models used cross-entropy loss function and were trained for 100 epochs with the initial learning rate of 0.005 decayed by a factor of 0.9 each epoch.

### Whole□Slide Inference

To classify whole-slide images, we aggregated patch-level prediction outputs. For each whole-slide image, we pre-processed the image by removing the white background and color markers. To aggregate the patch-level predictions, the ratio of patches from each class to the total number of patches from a slide is computed per whole-slide image. Bladder cancer is progressive, and there are mixed types of cancer cells in many whole-slides. Therefore, there are some low-risk patches in high-risk whole-slide images. We used a Gaussian process classifier for whole-slide inferencing. The classifier is trained on the patch ratios of whole-slide images from the training set and evaluated on the same validation set used in the patch-level analysis. Patch ratios of each classifier are given as input to the whole-slide inference classifier. Low-risk images usually have a higher ratio of patches labeled as PUNLMP and low-grade. High-risk images typically have a higher ratio of high-grade and invasive patches. Figure 2 shows patch ratios of various classes in four sample whole-slide images from different classes.

**Figure 2.**
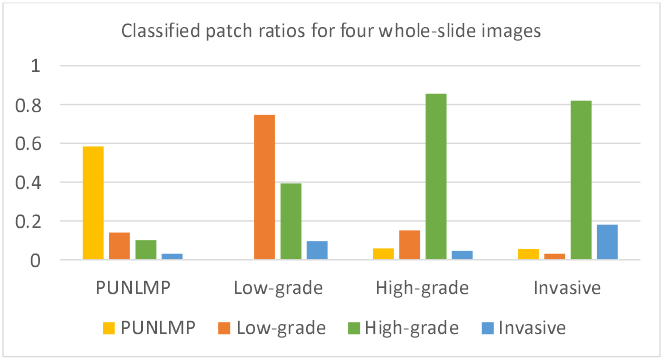
The ratio of patches in four whole-slide images. From left to right, these whole-slide images are with PUNLMP, low-grade, high-grade, and Invasive labels. The y-axis represents the ratio of the classified patches from each image.

We integrated the output of four binary patch-level classifiers to keep high-confidence patches and exclude low-confidence and normal patches in the whole-slide inference step. This process in our proposed pipeline is shown in Figure 3. If a patch is assigned to more than one of the labels, it indicates that the patch class label is unreliable and should be eliminated from the inference process. If all classifiers assign the label “others” to a patch, the patch is also eliminated. Our proposed inference method does not require hyper-parameter tuning as it does not rely on a threshold to eliminate low-confidence patches.

**Figure 3.**
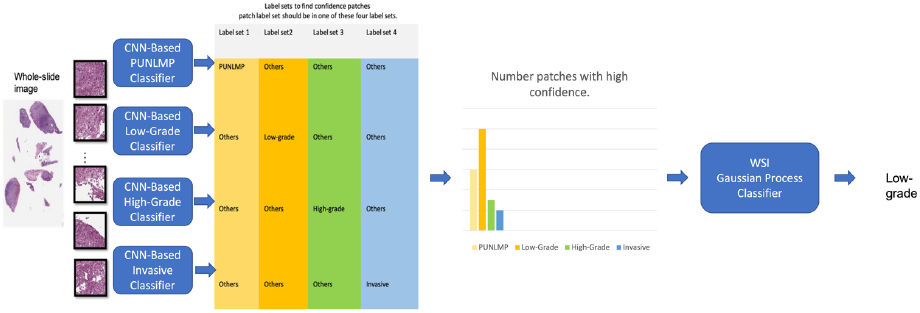
Patch extraction from whole-slide images. Each patch label set should be in one of the four presented combinations. Other patch label combinations where a patch can belong to more than one class indicate a low confident patch and are eliminated. This particular case corresponds to low-grade risk.

### Patient Survival Prediction

We analyzed the survival time of patients for low- and high-risk classes. Survival time was calculated from the date of diagnosis to the date of death for patients who did not survive or to the date when the Death Master File was queried for patients who survived [27]. We generated Kaplan-Meier survival curves for patients predicted as high-risk versus those predicted as low-risk. A Log-rank test was used to compare the survival between two predicted groups, considering the follow-up time. We used the Cox proportional hazards model [30] to estimate the effect size of our predicted risk group on patient survival.

### Evaluation Metrics and Statistical Analysis

To measure the efficacy and generalizability of our approach, we evaluated our trained model on 182 independent whole-slide images (WSIs) from the NHBCS dataset and 378 WSIs from the TCGA dataset. We used precision, recall, and the F1-score as evaluation metrics. The confusion matrix is also shown for error analysis. In addition, 95% confidence intervals were computed using the bootstrapping method with 10,000 iterations for all the metrics.

## Results

### Classification of Low-Risk and High-Risk Groups

Table 2 summarizes our model’s per-class and average evaluation metrics and the associated 95% CI for detecting low- and high-risk groups based on whole-slide images in the NHBCS test set. Our model achieved a weighted mean accuracy of 0.91, weighted mean precision of 0.91, weighted mean recall of 0.91, and weighted mean F1-score of 0.91 on the NHBCS test set.

**Table 2.**
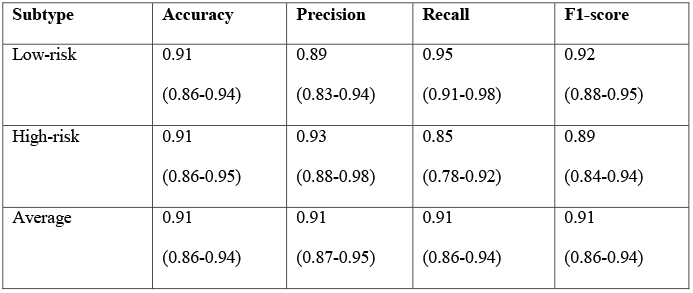
Model’s performance on 182 whole-slide images in the internal test set from NHBCS. The 95% confidence interval is also included for each measure.

| Subtype | Accuracy | Precision | Recall | F1-score |
| --- | --- | --- | --- | --- |
| Low-risk | 0.91<br>(0.86-0.94) | 0.89<br>(0.83-0.94) | 0.95<br>(0.91-0.98) | 0.92<br>(0.88-0.95) |
| High-risk | 0.91<br>(0.86-0.95) | 0.93<br>(0.88-0.98) | 0.85<br>(0.78-0.92) | 0.89<br>(0.84-0.94) |
| Average | 0.91<br>(0.86-0.94) | 0.91<br>(0.87-0.95) | 0.91<br>(0.86-0.94) | 0.91<br>(0.86-0.94) |

Table 3 shows the performance summary of our model on whole-slide images in the TCGA database under the study of urothelial bladder carcinoma. Of note, each case in the TCGA dataset may have more than one whole-slide image. Therefore, the images for these patients are aggregated in our study. The cancer stage of all TCGA images was T2 and above, i.e., high-risk, based on the patient metadata in the TCGA dataset. Although most patients in the TCGA cohort were in the high-risk group, high-risk histological patterns were absent on histology slides of a few patients based on the evaluation of our pathologist expert (B.R.). This is likely because only selected slides of each cases were uploaded to the TCGA database, and the selected slides may not represent the entire tumor. Therefore, based on the tumor morphology of WSIs available for these cases, we considered these cases as low-risk. On the TCGA dataset, our model achieved a weighted mean accuracy of 0.99, weighted mean precision of 0.99, weighted mean recall of 0.99, and weighted mean F1-score of 0.99. The confusion matrices for our model on the NHSBC and TCGA test sets are shown in Figure 4.

**Table 3.** Model’s performance on 378 whole-slide images from external TCGA dataset. The 95% confidence interval is also included for each measure.

| Subtype | Accuracy | Recall | Precision | F1-score |
| --- | --- | --- | --- | --- |
| Low-risk | 0.99<br>(0.98-1.00) | 1.0<br>(1.0-1.0) | 0.73<br>(0.46-0.99) | 0.84<br>(0.63-1.00) |
| High-risk | 0.99<br>(0.98-1.00) | 0.99<br>(0.98-1.0) | 1.0<br>(1.0-1.0) | 0.99<br>(0.98-1.00) |
| Average | 0.99<br>(0.98-1.00) | 0.99<br>(0.98-1.0) | 0.99<br>(0.98-1.00) | 0.99<br>(0.97-1.00) |

**Figure 4.**
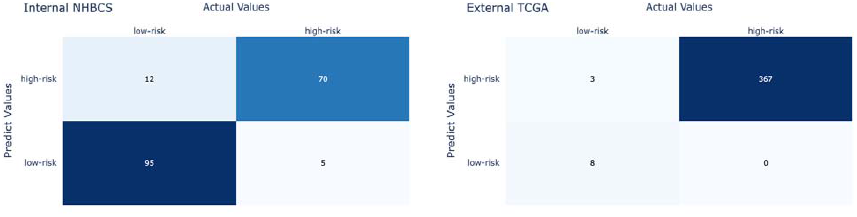
Each confusion matrix summarizes the model results compared to ground truth labels from pathologists on: (left) internal NHBCS test set and (right) external TCGA dataset.

### Prediction of Patient Survival

Figure 5 shows the Kaplan-Meier survival curve for patients from the internal NHBCS test set. The hazard ratio of overall survival using the predicted risk group by our model versus the tumor grade-defined risk groups for these patients is shown in Table 4. Figure 6 shows the Kaplan-Meier survival curve of patients from the external TCGA test set.

**Figure 5.**
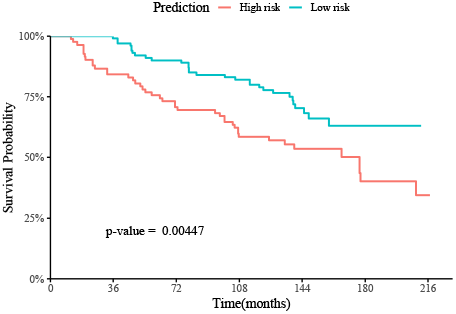
Kaplan-Meier survival curve of patients from the internal NHBCS test set with up to 216 months of follow-ups.

**Table 4.**
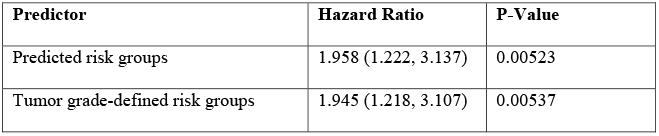
The hazard ratio of overall survival using the predicted risk group (predicted groups) versus the tumor grade-defined risk groups on patients from the internal NHBCS test set and the associated 95% CIs and P-values.

| Predictor | Hazard Ratio | P-Value |
| --- | --- | --- |
| Predicted risk groups | 1.958 (1.222, 3.137) | 0.00523 |
| Tumor grade-defined risk groups | 1.945 (1.218, 3.107) | 0.00537 |

**Figure 6.**
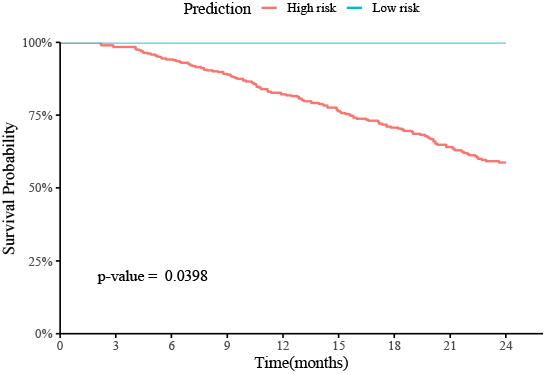
Kaplan-Meier survival curve of patients from the TCGA bladder cancer dataset with up to 48 months of follow-ups.

For the internal NHBCS test set, patients predicted as high-risk had worse overall survival than those predicted as low-risk, with a Log-rank p-value of 0.004 (Figure 5). The TCGA test patients were followed up to 216 months after the initial diagnosis, with a mean follow-up time of 123.8 months. The medium survival time of predicted high-risk patients was 177 months, while greater than 50% of the predicted low-risk patients survived until the end of their follow-ups. In the univariate Cox proportional hazards analysis using our predicted risk groups, the predicted high-risk group had an estimated hazard ratio of 1.958 (95% CI: 1.222–3.137, P-value=0.005) compared to the predicted low-risk group. This hazard ratio was slightly higher than the hazard ratio using the labels defined by the WHO/ISUP grading (Table 4).

Among 378 patients from the TCGA dataset, 367 were predicted as high-risk, and 11 as low-risk. Due to the small number of low-risk patients and their limited follow-up time of the TCGA data, we limited our survival analysis to the first 24 months after the initial diagnosis. There was no death event reported during the follow-up of the low-risk group and 35 death events reported in the high-risk group in the first 24 month of follow-up, with a Log-rank P-value of 0.04 (Figure 6). Of note, due to no events in the low-risk group, we could not estimate the hazard ratio and did not conduct Cox proportional hazards analysis on the TCGA dataset.

For the NHBCS test set, patients predicted as high-risk had worse overall survival compared to those predicted as low-risk, with a Log-rank P-value of 0.004 (Figure 5). The patients in the NHBCS dataset were followed up to 216 months after the initial diagnosis, with a mean follow-up time of 123.8 months. The median survival time of predicted high-risk patients was 177 months, while greater than 50% of the predicted low-risk patients survived until the end of their follow-ups.

## Discussion

The WHO has updated its bladder cancer grading guidelines several times since 1973 to align them more closely with disease recurrence and progression [3]. Based on the most recent update from WHO in 2016, PUNLMP, low-grade and high-grade with stage T1 bladder cancers are categorized as NMIBC, and high-grade cases with stage T2 and above are classified as MIBC. Detection and classification of bladder cancer histologic patterns under the microscope is critical for accurate prognosis and the appropriate treatment of patients; however, this histopathological assessment is a time-consuming and challenging task and suffers from a high variability rate among pathologists. Therefore, patients with NMIBC can incorrectly be diagnosed as high-grade cases, which might result in unnecessary treatment or even surgery which can affect patients’ quality of life [13] [31] [32]. In this study, we developed and evaluated a deep learning model to classify patients as high- or low-risk based on their whole-slide images to inform their prognosis and treatment. Our evaluation results on both internal and external datasets showed that this approach could potentially assist pathologists in their histopathological assessment, improve their accuracy and efficiency of diagnosis, and ultimately improve patient health outcomes.

As part of this study, we investigated developing a multi-class CNN model with four labels, including PUNLMP, low-grade, high-grade, and invasive. Although this model achieved a reasonable performance at the patch-level (See Table S1 in Supplemental Material), some classes, such as PUNLMP and invasive, achieved sub-optimal results. This outcome indicates that a single multi-class model cannot effectively handle the complexities of this task and achieve a good performance and generalization for all four classes. Notably, differentiating between PUNLMP and low-grade types has the lowest accuracy rate among clinicians due to their morphological similarities [13]. In addition, high-grade bladder cancer cells are found in any stage of the disease. Therefore, in our study, we used the ResNet-18 architecture as a backbone for binary classifications to differentiate between various classes instead of a single multi-class model. Each of our four binary CNN-based patch-level classifiers focuses on one class and differentiates that class from other subtypes. Our patch-level classification results (Table S1) indicate the high performance of our approach for this patch-level classification, as all binary classifiers achieved an F1-score of more than 0.79.

As the primary whole-slide level classification outcomes, we focused on identifying low- and high-risk groups for bladder cancer based on histology slides, where low-risk class includes PUNLMP and low-grade cases and the high-risk class includes high-grade and invasive cases. The differentiation between these two risk groups has a significant clinical impact on patient prognosis and treatment. For whole-slide inferencing, we built a Gaussian process classifier based on the distribution of the classified patches from each slide.

To demonstrate the generalizability of our model, in addition to evaluating it on 182 whole-slide images in our internal test set from NHBCS, we also evaluated our approach on 378 whole-slide images from TCGA as an external test set. Our approach achieved the weighted average F1-score of 0.91 (95% CI: 0.86–0.94) on the internal NHBCS test set. Our model achieved the weighted average F1-score of 0.99 (95% CI: 0.97–1.00) on the external TCGA test set. Of note, TCGA metadata information showed that all the cases in this dataset belong to the high-risk class. That said, a few cases were identified by our study’s expert pathologist (B.R.) as low risk based on their histology images, likely due to the selected slides included in the TCGA dataset may not represent the entire tumor.

Finally, we computed Kaplan-Meier survival curves for patients predicted as high-risk versus those predicted as low-risk for both NHBCS and TCGA test sets. Patients predicted as high-risk had worse overall survival than those predicted as low-risk, with Log-rank P-values of 0.004 and 0.039 on the NHBCS and TCGA test sets, respectively. Also, our predicted high-risk group in the NHBCS test had an estimated hazard ratio of 1.958 (95% CI: 1.222–3.137, P-value=0.005) compared to the predicted low-risk group, which was slightly higher than the hazard ratio using the labels defined by the WHO/ISUP grading.

In future work, we plan to expand our model to distinguish between high-grade invasive and high-grade noninvasive classes, which is clinically helpful to determine the progression and reoccurrence of bladder cancer. Because we had a limited number of muscle-invasive cases in our datasets, building such a model for this differentiation was not feasible in the current study. We plan to collect additional data and develop new data augmentation techniques, such as generative adversarial networks (GANs), to tackle the dataset imbalance. Such techniques can mitigate the effects of unbalanced data by preventing overfitting and thus improving overall performance [33]. In addition, we consider including vision transformers in our future pipeline to improve our high-resolution image encoding approach [34]. As future work, we also plan to deploy the developed model for histopathological characterization of whole-slide images for bladder cancer as a computer-aided diagnosis system in clinical settings. We plan to conduct a prospective study to validate our approach in clinical practice and evaluate its impact on health outcomes.

## Data Availability

All data produced in the present study are available upon reasonable request to the authors

## Acknowledgements

This research was supported in part by grants from the US National Library of Medicine (R01LM012837), the US National Cancer Institute (R01CA249758), and the US National Institute of General Medical Sciences (P20GM104416).

## References

[1] K. Saginala, A. Barsouk, J. S. Aluru, P. Rawla, S. A. Padala and A. Barsouk, “Epidemiology of Bladder Cancer,” Medical sciences, vol. 8, no. 1, p. 15, 13 March 2022.

[2] A. Anastasiadis and T. M. de Reijke, “Best practice in the treatment of nonmuscle invasive bladder cancer,” Therapeutic advances in urology, vol. 4, pp. 13–32, 2012.

[3] R. Engers, “Reproducibility and reliability of tumor grading in urological neoplasms,” World journal of urology, vol. 25, pp. 595–605, 2007.

[4] V. Soukup, O. vCapoun, D. Cohen, V. Hernandez, M. Babjuk, M. Burger, E. Comperat, P. Gontero, T. Lam and S. MacLennan, “Prognostic performance and reproducibility of the 1973 and 2004/2016 World Health Organization Grading classification systems in non--muscle-invasive bladder cancer: a European Association of Urology non-muscle invasive bladder cancer guidelines panel sys,” European urology, vol. 72, pp. 801–813, 2017.

[5] A. E. Hentschel, B. W. van Rhijn, J. Brundl, E. M. Comperat, K. Plass, O. Rodriguez, J. D. S. Henriquez, Hern, V. ez, E. de la Pena and I. Alemany, “Papillary urothelial neoplasm of low malignant potential (PUN-LMP): Still a meaningful histo-pathological grade category for Ta, noninvasive bladder tumors in 2019?,” in Urologic Oncology: Seminars and Original Investigations, Elsevier, 2020, pp. 440–448.

[6] W. M. Murphy, K. Takezawa and N. A. Maruniak, “Interobserver discrepancy using the 1998 World Health Organization/International Society of Urologic Pathology classification of urothelial neoplasms: practical choices for patient care,” The Journal of urology, vol. 168, pp. 968–972, 2002.

[7] J. K. Kim, K. C. Moon, C. W. Jeong, C. Kwak, H. H. Kim and J. H. Ku, “Papillary urothelial neoplasm of low malignant potential (PUNLMP) after initial TUR-BT: comparative analyses with noninvasive low-grade papillary urothelial carcinoma (LGPUC),” Journal of Cancer, vol. 8, p. 2885, 2017.

[8] E. M. Comperat, M. Burger, P. Gontero, A. H. Mostafid, J. Palou, M. Roupret, B. W. van Rhijn, S. F. Shariat, R. J. Sylvester and R. Zigeuner, “Grading of urothelial carcinoma and the new “World Health Organisation classification of tumours of the urinary system and male genital organs 2016”,” European urology focus, vol. 5, pp. 457–466, 2019.

[9] A. Robertson, J. S. Beck, R. Burnett, S. Howatson, F. Lee, A. Lessells, K. McLaren, S. Moss, J. Simpson and G. Smith, “Observer variability in histopathological reporting of transitional cell carcinoma and epithelial dysplasia in bladders.,” Journal of clinical pathology, vol. 43, pp. 17–21, 1990.

[10] K. Yorukoglu, B. Tuna, E. Dikicioglu, E. Duzcan, A. Isisag, S. Sen, U. Mungan and Z. Kirkali, “Reproducibility of the 1998 World Health Organization/International Society of Urologic Pathology classification of papillary urothelial neoplasms of the urinary bladder,” Virchows Archiv, vol. 443, pp. 734–740, 2003.

[11] H. W. Herr, S. M. Donat and V. E. Reuter, “Management of Low Grade Papillary Bladder Tumors,” The Journal of Urology, vol. 178, no. 4, pp. 1201–1205, 2007.

[12] J. I. Epstein, M. B. Amin, V. R. Reuter, F. K. Mostofi and B. C. Conference Committee, “The World Health Organization International Society of Urological Pathology consensus classification of urothelial (transitional cell) neoplasms of the urinary bladder,” The American journal of surgical pathology, vol. 22, pp. 1435–1448, 1998.

[13] W. M. Murphy, K. Takezawa and N. A. Maruniak, “Interobserver discrepancy using the 1998 World Health Organization,” The Journal of urology, vol. 168, pp. 968--972, 2002.

[14] K.-L. Hua, C.-H. Hsu, S. C. Hidayati, W.-H. Cheng and Y.-J. Chen, “Computer-aided classification of lung nodules on computed tomography images via deep learning technique,” OncoTargets and therapy, vol. 8, 2015.

[15] S. Tabibu, P. Vinod and C. Jawahar, “Pan-Renal Cell Carcinoma classification and survival prediction from histopathology images using deep learning,” Scientific reports, vol. 9, pp. 1 –9, 2019.

[16] N. Tomita, Y. Y. Cheung and S. Hassanpour, “Deep neural networks for automatic detection of osteoporotic vertebral fractures on CT scans,” Computers in biology and medicine, vol. 98, pp. 8–15, 2018.

[17] T. Araujo, G. Aresta, E. Castro, J. Rouco, P. Aguiar, C. Eloy, A. Polonia and A. Campilho, “Classification of breast cancer histology images using convolutional neural networks,” PloS one, p. e0177544, 2017.

[18] N. Coudray, P. S. Ocampo, T. Sakellaropoulos, N. Narula, M. Snuderl, D. Fenyo, A. L. Moreira, N. Razavian and A. Tsirigos, “Classification and mutation prediction from non-small cell lung cancer histopathology images using deep learning,” Nature medicine, vol. 24, pp. 1559–1567, 2018.

[19] A. Cruz-Roa, H. Gilmore, A. Basavanhally, M. Feldman, S. Ganesan, N. N. Shih, J. Tomaszewski, F. A. Gonzalez and A. Madabhushi, “Accurate and reproducible invasive breast cancer detection in whole-slide images: A Deep Learning approach for quantifying tumor extent,” Scientific reports, vol. 7, pp. 1–14, 2017.

[20] S. Jiang, G. J. Zanazzi and S. Hassanpour, “Predicting Prognosis and IDH Mutation Status for Patients with Lower-Grade Gliomas Using Whole Slide Images”,” Scientific Reports, vol. 11, no. 1, p. 16849, 2021.

[21] J. W. Wei, A. A. Suriawinata, L. J. Vaickus, B. Ren, X. Liu, M. Lisovsky, N. Tomita, B. Abdollahi, A. S. Kim, D. C. Snover, J. A. Baron, E. L. Barry and S. Hassanpour, “Evaluation of a Deep Neural Network for Automated Classification of Colorectal Polyps on Histopathologic Slides,” JAMA Network Open, vol. 3, no. 4, pp. e203398–e203398, 2020.

[22] M. R. Karagas, T. D. Tosteson, J. Blum, J. S. Morris, J. A. Baron and B. Klaue, “Design of an epidemiologic study of drinking water arsenic exposure and skin and bladder cancer risk in a US population.,” Environmental health perspectives, vol. 106, pp. 1047–1050, 1998.

[23] K. Kelsey, T. Hirao, A. Schned, S. Hirao, T. Devi-Ashok, H. Nelson, A. Andrew and M. Karagas, “A population-based study of immunohistochemical detection of p53 alteration in bladder cancer,” British journal of cancer, vol. 90, pp. 1572–1576, 2004.

[24] E. F. Sverrisson, M. S. Zens, D. L. Fei, A. Andrews, A. Schned, D. Robbins, K. T. Kelsey, H. Li, J. DiRenzo and M. R. Karagas, “Clinicopathological correlates of Gli1 expression in a population-based cohort of patients with newly diagnosed bladder cancer,” in Urologic Oncology: Seminars and Original Investigations, Elsevier, 2014, pp. 539–545.

[25] J. S. Colt, M. R. Karagas, M. Schwenn, D. Baris, A. Johnson, P. Stewart, C. Verrill, L. E. Moore, J. Lubin and M. H. Ward, “Occupation and bladder cancer in a population-based case-control study in Northern New England,” Occupational and environmental medicine, vol. 68, pp. 239–249, 2011.

[26] cancer.gov, “The Cancer Genome Atlas,” [Online]. Available: https://www.cancer.gov/tcga. x[Accessed 3 September 2021].

[27] Q. Song, J. Seigne, A. Schned, K. Kelsey, M. Karagas and S. Hassanpour, “A machine learning approach for long-term prognosis of bladder cancer based on clinical and molecular features,” AMIA Jt Summits Transl Sci Proc, pp. 607-616. PMID: 32477683; PMCID: PMC7233061., 20 May 2020.

[28] Computation Pathology Group at Radboud University Medical Center, “Automated Slide Analysis Platform (ASAP),” [Online]. Available: https://computationalpathologygroup.github.io/. x[Accessed 5 September 2021].

[29] K. He, X. Zhang, S. Ren and J. Sun, “Deep residual learning for image recognition,” 2016 IEEE Conference on Computer Vision and Pattern Recognition (CVPR), 2016.

[30] D. R. Cox, “Regression Models and Life-Tables,” Journal of the Royal Statistical Society, Series B., vol. 34, no. 2, p. 87–220, 1972.

[31] R. Veeratterapillay, R. Heer, M. I. Johnson, R. Persad and C. Bach, “High-risk non-muscle-invasive bladder cancer—therapy options during intravesical BCG shortage,” Current urology reports, vol. 17, pp. 1–7, 2016.

[32] N. E. Mohamed, P. C. Herrera, S. Hudson, T. A. Revenson, C. T. Lee, D. Z. Quale, C. Zarcadoolas, S. J. Hall and M. A. Diefenbach, “Muscle invasive bladder cancer: examining survivor burden and unmet needs,” The Journal of urology, vol. 191, pp. 48–53, 2014.

[33] J. Wei, A. Suriawinata, L. Vaickus, B. Ren, X. Liu, J. Wei and S. Hassanpour, “Generative Image Translation for Data Augmentation in Colorectal Histopathology Images,” 2019.

[34] P. Zhang, X. Dai, J. Yang, B. Xiao, L. Yuan, L. Zhang and J. Gao, “Multi-Scale Vision Longformer: A New Vision Transformer for High-Resolution Image Encoding,” Proceedings of the IEEE/CVF International Conference on Computer Vision (ICCV), pp. 2998–3008, October 2021.

[35] P. Khosravi, E. Kazemi, M. Imielinski, O. Elemento and I. Hajirasouliha, “Deep convolutional neural networks enable discrimination of heterogeneous digital pathology images,” EBioMedicine, vol. 27, pp. 317–328, 2018.

[36] R. R. Selvaraju, M. Cogswell, A. Das, R. Vedantam, D. Parikh and D. Batra, “Grad-cam: Visual explanations from deep networks via gradient-based localization,” in Proceedings of the IEEE international conference on computer vision, 2017, pp. 618–626.

